# Research Letter: Genetically determined blood pressure traits are associated with increased risk of aortic dissection: a Mendelian randomization study

**DOI:** 10.1101/2022.02.06.22270435

**Authors:** Jiawei Zhou, Jianfeng Lin, Yuehong Zheng

## Abstract

**Objective:** To identify blood pressure traits influencing aortic dissection susceptibility using Mendelian randomization (MR).

**Methods:** Summary statistics of AD genome-wide association studies (GWAS) was obtained from FinnGen R5 release, which included 470 individuals with AD and 206541 controls. Firstly, inverse variance weighted (IVW) was used to calculate the overall OR for blood pressure traits. Further, directional pleiotropy and heterogeneity were tested as well as MR pleiotropy residual sum and outlier (MR-PRESSO) method.

**Results:** Positive associations were observed between DBP (OR=1.13, 95%CI 1.09-1.18, P-adjusted < 0.001), hypertension (OR=9.71, 95%CI 2.45-38.25, P-adjusted = 0.006) and AD. Further, there was potential causal relationship between MAP and AD (OR=1.15, 95%CI 1.02-1.30, P-unadjusted = 0.020). No significant association between SBP or PP and AD in the European populations were observed. No directional pleiotropy was observed. Heterogeneity was only detected in the genetic instruments for SBP (Cochran’s Q statistics = 491.7, P = 0.010). The MR-PRESSO suggested one outlier SNP for SBP and one for PP which did not affect the result above.

**Conclusion:** In this study, we identified a forward causation between hypertension, SBP and AD. In addition, there were nominal significant causations between MAP and AD. No significant causal relationships were detected between other investigated blood pressure traits (SBP, PP) and AD.

---

A recent meta□analysis enrolled 8 cohort studies with a total of 2,818 cases and 4,563,501 participants to investigate the association between hypertension & blood pressure elevation and aortic dissection (AD).(1) These findings were of great interest as discussed below. The summary relative risks (RRs) for hypertension were 3.07 (95% CI 2.15-4.38, I^2^=76.7%). Furthermore, the RRs were 1.39 (95% CI: 1.16-1.66, I^2^=47.7%) and 1.79 (95% CI: 1.51-2.12, I^2^=57.0%) for per increase of 20 mmHg in systolic blood pressure (SBP) and per increase of 10 mmHg in diastolic blood pressure (DBP), respectively. The results indicated positive dose-related associations between elevated SBP, DBP and AD in which the effect of DBP on AD was even stronger. Due to potential confounders and reverse causation bias, observational studies could not investigate causality between hypertension and AD. With Two-Sample Mendelian randomization (MR) analysis, the influence of confounding and reverse causality could be avoided to estimate the causal effect of a risk factor (exposure) on an outcome.

In this MR analysis, we obtained summary statistics of AD genome-wide association studies (GWAS) from FinnGen R5 release, which included 470 individuals with AD and 206541 controls, with a total of 16,380,465 genotyped SNPs. To construct the genetic instruments for each blood pressure trait, we used GWASs conducted in large cohorts of European. Firstly, we included single-nucleotide polymorphisms (SNPs) associated with each trait at the genome-wide significance level (P<5×10^−8^), and selected the SNP with the strongest association when linkage disequilibrium was present (r^2^>0.1). Secondly, we excluded SNPs that were unavailable in the AD GWAS. Finally, the SNPs used for genetic instruments were 196, 422, 427, 54, and 11 for hypertension (2), SBP (3), DBP (3), pulse pressure (PP) (3), and mean arterial pressure (MAP) (4), respectively. We used inverse variance weighted (IVW) was used to calculate the overall OR. We also performed MR-Egger (MRE) regression to assess directional pleiotropy and calculated Cochran’s Q statistics to test heterogeneity test. MR pleiotropy residual sum and outlier (MR-PRESSO) method were further used to exclude potentially horizontally pleiotropic SNPs (P<0.10). Benjamini–Hochberg method was used to calculate multiple testing correction of P-value. P value smaller than 0.05 was considered as statistically significant. All statistical analyses were performed using R 4.0.5.

The corresponding F-statistics of genetic instrument for each blood pressure trait were 29.8-462.7, 29.7-627.5, 29.7-815.8, 36.6-134.2, 30.5-83.3, respectively, suggesting that the instruments were unlikely to suffer from weak instrument bias. The variance explained ranged from 0.4% for PP to 4.5% for DBP. Our MR study identified, for the first time, the causality between blood pressure traits and AD with the method of Mendelian Randomization (Figure 1). The strongest positive associations were observed between DBP (OR=1.13, 95%CI 1.09-1.18, P-adjusted < 0.001), hypertension (OR=9.71, 95%CI 2.45-38.25, P-adjusted = 0.006) and AD. Moreover, there was potential causal relationship between MAP and AD (OR=1.15, 95%CI 1.02-1.30, P-unadjusted = 0.020). Nevertheless, the study failed to reveal the significant association between SBP or PP and AD in the European populations. We did not find any directional pleiotropy, and only observed heterogeneity in the genetic instruments for SBP (Cochran’s Q statistics = 491.7, P = 0.010). The MR-PRESSO suggested one outlier SNP for SBP and one for PP. After excluding the outlier SNPs, the estimates were only subtly changed (SBP: OR=1.01, 95%CI 0.99-1.04, P-adjusted = 1; PP: OR=0.92, 95%CI 0.77-1.10, P-adjusted = 1), while heterogeneity for SBP-related SNPs was diminished (Cochran’s Q statistics = 448.8, P = 0.160).

**Figure 1.**
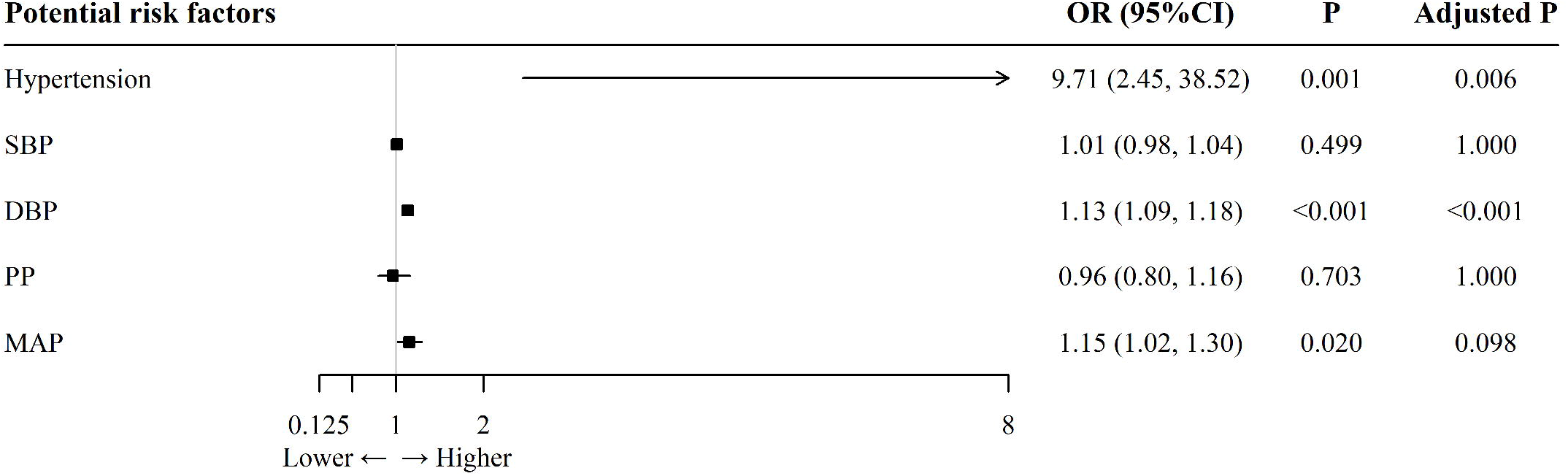
The association between modifiable risk factors and aortic dissection using the inverse-variance weighted method.

Based on this, our MR study proved the positive causality between hypertension, DBP and AD which were consistent with present epidemiological evidence.(1) However, the conclusion of causal relationship between SBP and AD was not supported by existing literature. The different roles for SBP & DBP were also observed in other cardiovascular disease awaiting further investigation.(5)

In conclusion, this study identified a forward causation between hypertension, SBP and AD for the first time. In addition, there were nominal significant causations between MAP and AD. No significant causal relationships were detected between other investigated blood pressure traits (SBP, PP) and AD. For the validated exposure, detailed mechanisms underlying these causations need to be investigated in future studies.

## Data Availability

All data produced in the present study are available upon reasonable request to the authors.

## Acknowledgments

None.

## Authors’ contributions

YZ is responsible for substantial contributions to the conception and substantively revision. JZ is responsible for study design, data acquisition, manuscript writing, and substantively revision. JL is responsible for study design, data analysis, data interpretation, manuscript writing, and substantively revision. All the authors approved the submitted version and agreed both to be personally accountable for the author’s own contributions and to ensure that questions related to the accuracy or integrity of any part of the work.

## Funding

This work is supported by the Major Research Program of Natural Science Foundation of China (51890894), Natural Science Foundation of China (81770481 and 82070492), Chinese Academy of Medical Sciences, Innovation Fund for Medical Sciences (2021-I2M-C&T-A-006) and Innovation Fund for Health and Longevity in China (JC2021CL006). The funders had no role in study design, data collection and analysis, decision to publish, or preparation of the manuscript.

## Availability of data and materials

No new data were created or analyzed in this study. All the data used in this study could be downloaded from publicly available databases or published articles.

## Declarations

### Ethics approval and consent to participate

Not applicable.

### Consent for publication

Not applicable.

### Competing interests

The authors declare that they have no competing interests.

## Notes

### Competing Interest Statement

The authors have declared no competing interest.

